# Hit hardest, healed least: disruption and incomplete recovery of public mental-health outpatient services across two COVID-19 waves in India, 2017-2022

**DOI:** 10.64898/2026.09.11.26362835

**Authors:** Vartika Mishra, Koshtubh Singh Parihar

**Affiliations:** Department of Pathology, Ruxmaniben Deepchand Gardi Medical College, Ujjain, Madhya Pradesh, India; Government Medical College Datia, Datia, Madhya Pradesh, India

**Keywords:** COVID-19, mental health services, India, interrupted time series, outpatient care, health management information system, Tele-MANAS

## Abstract

**Background:** India’s COVID-19 waves disrupted essential health services, but the impact on public mental healthcare has been documented only through provider surveys and single-facility reports. We quantified the disruption and recovery of mental-health outpatient services nationally, using routine administrative data, and compared it against physical-health services over the same period.

**Methods:** Interrupted time-series analysis of India’s Health Management Information System (HMIS), April 2017 to March 2022. Monthly mental-illness outpatient (OPD) visit counts and adolescent counselling contacts were extracted for all states and districts. Expected counts from March 2020 were projected from log-linear trend plus calendar-month seasonality fitted on the 35 pre-pandemic months, with a no-trend same-month-mean counterfactual as sensitivity analysis. District-level recovery was classified from final-year (April 2021-March 2022) observed/expected ratios among districts with stable reporting.

**Results:** Public mental-illness OPD visits, which had grown from 4.1 to 6.1 million per year between 2017-18 and 2019-20, fell to 31% of expected in April 2020 — a deeper acute shock than any maternal-child health indicator in the same reporting system (38-84%). Recovery was slower and less complete: mental-illness OPD averaged 43% of expected during the Delta wave and 58% in the final study year, versus around 90% for a composite of physical-health services. The cumulative two-year deficit was 18.5-47.1% across counterfactuals; absolute visits in 2021-22 (5.36 million) remained below the 2019-20 actual (6.10 million). Among 220 districts with stable reporting, 79% had not returned to within 85% of expected by the final year, the median district operated at 41% of expected (IQR 16-78%), and districts hit hardest in April-June 2020 remained furthest behind (Spearman rho=0.60). Adolescent counselling contacts showed the same twin-shock, partial-recovery pattern (final-year ratio 72-74%). Cumulative state deficits exceeded 60% in eleven states.

**Conclusion:** Public mental-health outpatient care in India was hit harder by COVID-19 than physical-health services and, uniquely among major service lines, had not recovered two years on. These findings provide the missing quantitative baseline for evaluating India’s post-pandemic mental-health investments, including the National Tele Mental Health Programme (Tele-MANAS) launched in October 2022, and argue for district-targeted service restoration rather than uniform national programming.

## Introduction

The COVID-19 pandemic disrupted health services worldwide, and mental healthcare was among the most affected service lines globally: WHO pulse surveys found mental, neurological and substance-use services among the most frequently interrupted throughout 2020 [1]. In India — where the National Mental Health Survey had already documented a treatment gap of 70-92% for common mental disorders before the pandemic [2] — any interruption of the public mental-health delivery system falls on a population with few alternatives, since public facilities under the District Mental Health Programme (DMHP) are the only affordable source of psychiatric care for much of the population.

The Indian evidence on this disruption is, however, almost entirely indirect. A survey of Indian Psychiatric Society members in May 2020 documented steep declines in private psychiatric practice [3]; a survey of 109 psychiatric training centres described suspension of outpatient, inpatient and electroconvulsive therapy services during the lockdown [4]; and single-facility reports describe outpatient volumes collapsing and partial adaptation through teleconsultation [5]. What is missing is a quantification: how large was the disruption to public mental healthcare nationally, how did it compare with the better-studied disruption of physical-health services, and — most importantly for policy — did it recover?

India’s Health Management Information System (HMIS) records monthly mental-illness outpatient attendance from public facilities in every district, offering a direct, national, administrative measure. We analysed five years of these records spanning both major COVID-19 waves to quantify the disruption, benchmark it against physical-health services from the same reporting system, measure recovery through March 2022, and characterise district-level heterogeneity. The end of our study window coincides with the launch of the National Tele Mental Health Programme (Tele-MANAS, October 2022), for which these findings constitute the immediate pre-launch baseline [6].

## Methods

### Data source and indicators

We used monthly district-level HMIS reports, fiscal years 2017-18 to 2021-22 (April 2017-March 2022; 2,185 state-month report files parsed with zero extraction errors). The primary indicator was ‘Outpatient - Mental illness’ (item 14.1.5), the count of outpatient attendances for mental illness in public facilities. Secondary indicators were adolescent counselling contacts at Adolescent Friendly Health Clinics (items 12.1.3.a/b, girls and boys counselled). For comparison with physical-health services we used the observed/expected series for institutional deliveries, antenatal registrations and DPT3/Pentavalent-3 immunization from a companion analysis of the same archive using the identical model.

### Statistical analysis

For each indicator at national and state level, expected monthly counts from March 2020 onward were projected from ordinary least-squares models of log counts on linear time and calendar-month indicators, fitted to April 2017-February 2020. Disruption was summarised as observed/expected (O/E) ratios for the acute first wave (March-June 2020), the inter-wave period (July-December 2020), the Delta wave (April-June 2021), and the final year (July 2021-March 2022), with cumulative deficits over March 2020-March 2022. Because pre-pandemic mental-health OPD volumes were growing steeply (approximately 50% over three years), trend projection produces a generous counterfactual; all national deficits were therefore re-estimated against a no-trend counterfactual (same-calendar-month mean of the two complete pre-pandemic years) and results are reported as ranges. The same model was fitted per district (minimum 100 visits/month pre-pandemic mean); districts with an acute- or late-period O/E ratio above 3 were excluded as reporting discontinuities (84 of 304 fitted districts), and district scarring (association between acute shock and final-year recovery) was assessed with Spearman correlation, which is robust to residual outliers. Analyses used Python 3.13; reporting follows RECORD guidance [7].

## Results

### National disruption and the mental-physical gap

Public mental-illness OPD visits grew from 4.07 million (FY 2017-18) to 6.10 million (FY 2019-20), an increasing trend that the pre-pandemic model captured closely (Figure 1, top). In April 2020 visits fell to 175,056 against 486,869 a year earlier — 31% of model-expected — the deepest acute shock of any major service indicator in the same reporting system (institutional deliveries 84%, ANC registrations 59%, DPT3 immunization 39% of expected in the same month). Adolescent counselling contacts fell in parallel (30-32% of expected in April 2020; Table 1).

**Table 1.** National observed/expected ratios (%) by period and cumulative deficits (March 2020-March 2022) under two counterfactuals, mental-health service indicators.

| Indicator | Apr 2020 | Mar-Jun 2020 | Jul-Dec 2020 | Delta (Apr-Jun 2021) | Jul 2021-Mar 2022 | Deficit % (trend) | Deficit % (flat) |
| --- | --- | --- | --- | --- | --- | --- | --- |
| Mental-illness outpatient visits | 31 | 46 | 46 | 43 | 58 | -47 | -19 |
| Adolescent girls counselled | 30 | 45 | 49 | 60 | 72 | -40 | -22 |
| Adolescent boys counselled | 32 | 46 | 48 | 57 | 74 | -39 | -20 |

**Figure 1.**
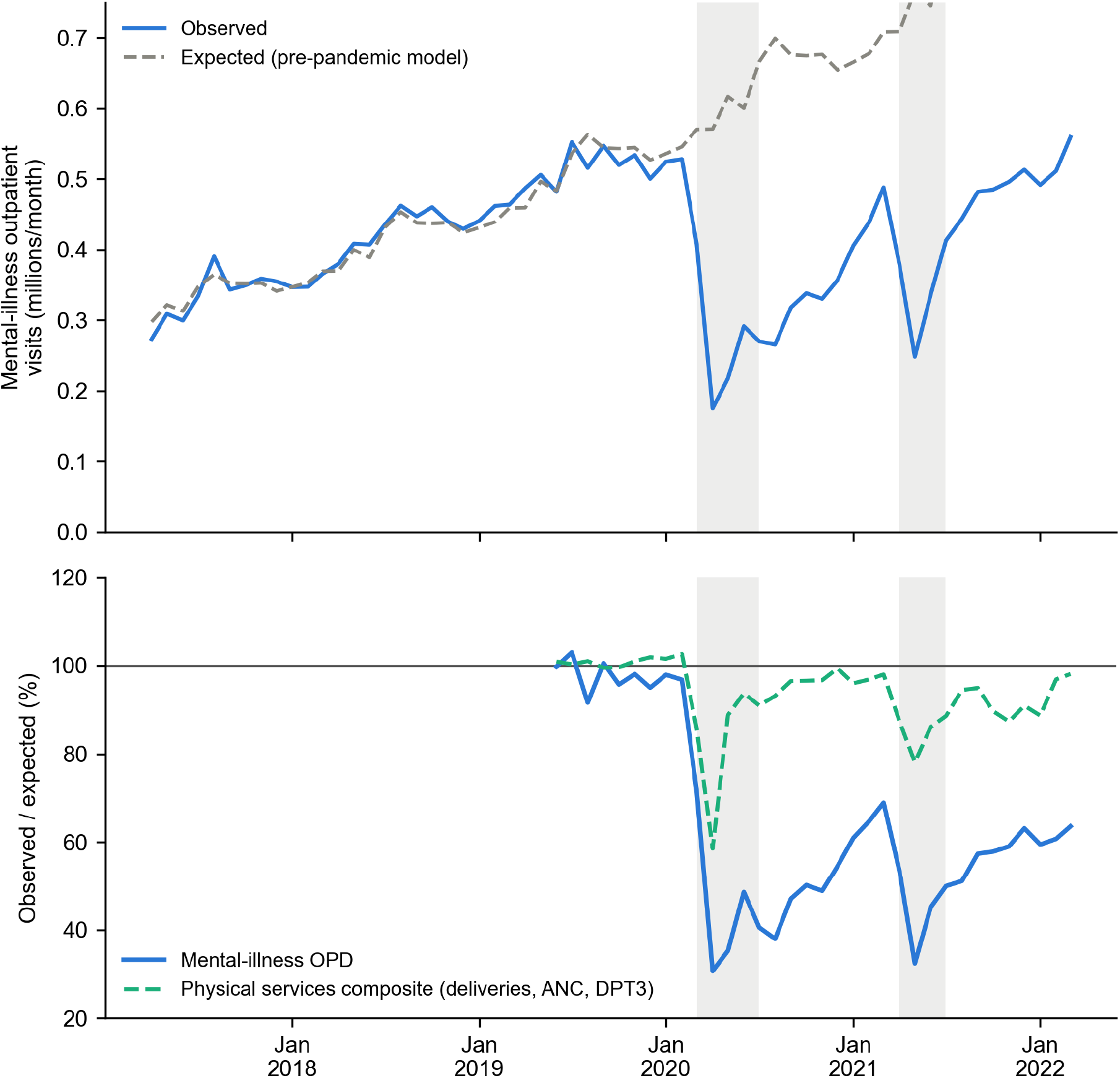
National mental-illness outpatient visits, observed versus expected (top), and observed/expected ratio versus a physical-services composite (bottom), 2017-2022. Shaded bands: first lockdown and Delta wave.

The distinctive feature of mental-health services was not the depth of the shock but the failure to recover (Figure 1, bottom). While immunization returned to expected levels within months and the physical-services composite recovered to roughly 90% of expected through 2021-22, mental-illness OPD plateaued at 46% of expected through late 2020, fell to 43% during the Delta wave, and reached only 58% of expected in the final study year. In absolute terms, FY 2021-22 volumes (5.36 million) remained 12% below the last pre-pandemic year’s actual volumes (6.10 million) — the only major HMIS service line still below its pre-pandemic absolute level two years after the pandemic began. The cumulative two-year deficit was 47.1% of expected under the trend counterfactual and 18.5% under the conservative no-trend counterfactual — between 2.2 and 8.4 million missed mental-health outpatient contacts. Adolescent counselling recovered somewhat further (final-year ratio 72-74%; cumulative deficit 20-40%).

### State and district heterogeneity

Cumulative deficits exceeded 60% of expected visits in eleven states, with the largest proportional losses in Jharkhand (-92%), Himachal Pradesh (-90%), Mizoram (-89%), Chandigarh (-82%), Punjab (-75%) and Uttar Pradesh (-73%); Kerala and Karnataka, which together account for a quarter of national public mental-health OPD volume, lost 60-63% (Table 2, Figure 2). Only Gujarat (-13%), Bihar (-21%) and Tamil Nadu (-23%) kept cumulative losses below a quarter of expected volume. Among 220 districts with stable reporting series, 79% remained below 85% of expected in the final year and 57% below half; the median district operated at 41% of expected (IQR 16-78%). Districts with the deepest April-June 2020 collapse were systematically the districts furthest from recovery in the final year (Spearman rho=0.60; Figure 3), the same ‘scarring’ pattern documented for physical services but at a much lower overall level of recovery.

**Table 2.** State cumulative mental-illness OPD deficits, March 2020-March 2022 (states with >100,000 expected visits; thousands of visits).

| State | Observed (000s) | Expected (000s) | Deficit (%) |
| --- | --- | --- | --- |
| Jharkhand | 31 | 377 | -91.7 |
| Himachal Pradesh | 63 | 602 | -89.5 |
| Mizoram | 19 | 176 | -89.3 |
| Chandigarh | 81 | 441 | -81.6 |
| Punjab | 428 | 1,724 | -75.2 |
| Uttar Pradesh | 511 | 1,876 | -72.7 |
| Haryana | 239 | 698 | -65.8 |
| Chhattisgarh | 178 | 499 | -64.3 |
| Kerala | 724 | 1,949 | -62.8 |
| Assam | 103 | 271 | -62.1 |
| Karnataka | 829 | 2,091 | -60.3 |
| Madhya Pradesh | 248 | 617 | -59.8 |
| Maharashtra | 457 | 1,100 | -58.5 |
| Rajasthan | 811 | 1,769 | -54.2 |
| Jammu & Kashmir | 119 | 245 | -51.5 |
| Odisha | 195 | 374 | -47.8 |
| Andhra Pradesh | 420 | 624 | -32.8 |
| Delhi | 336 | 480 | -29.9 |
| West Bengal | 1,187 | 1,650 | -28.1 |
| Tamil Nadu | 1,739 | 2,249 | -22.7 |
| Bihar | 85 | 107 | -20.8 |
| Gujarat | 580 | 669 | -13.3 |

**Figure 2.**
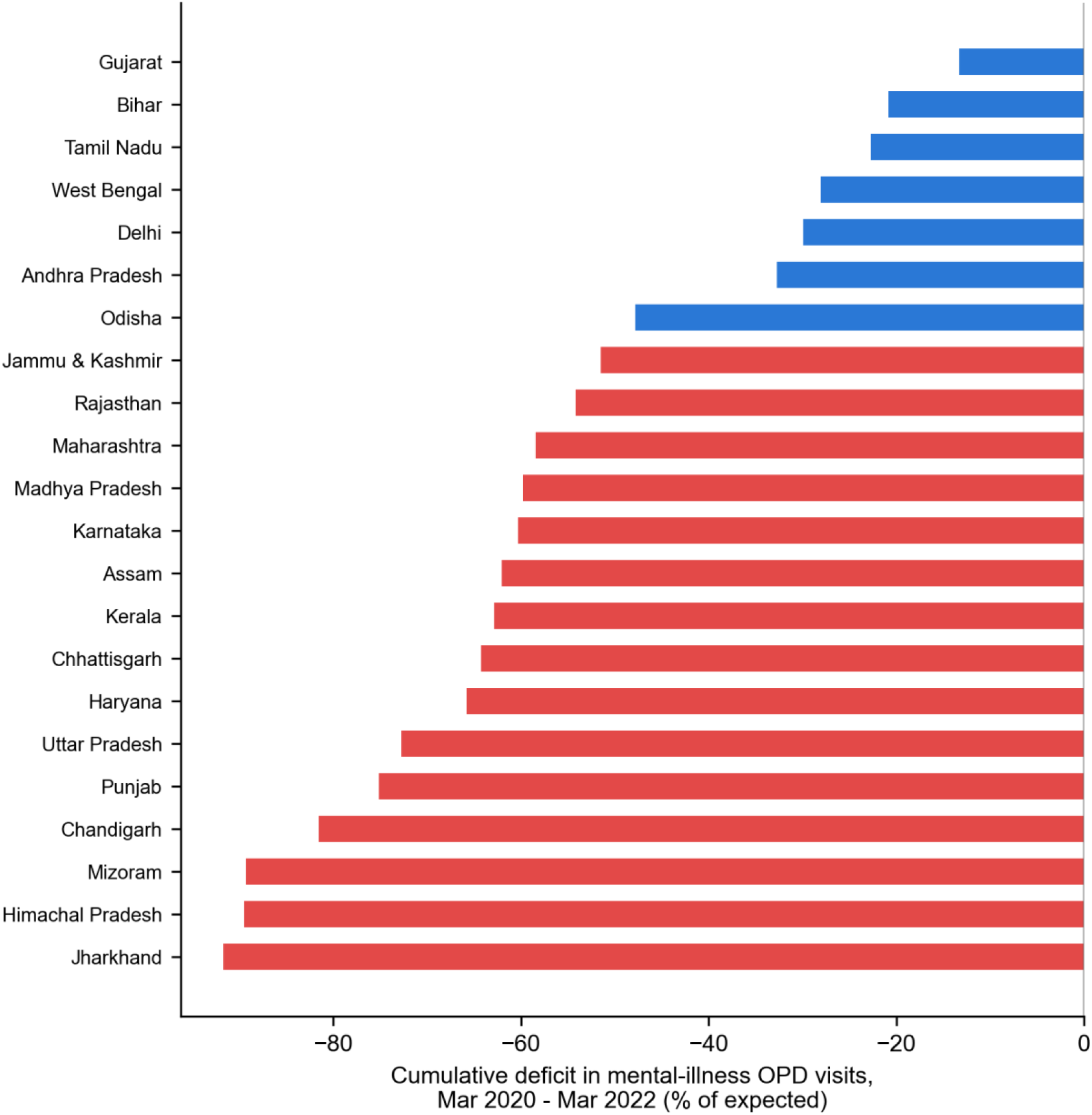
State cumulative mental-illness OPD deficit, March 2020-March 2022 (% of expected; red: deficit exceeding 50%).

**Figure 3.**
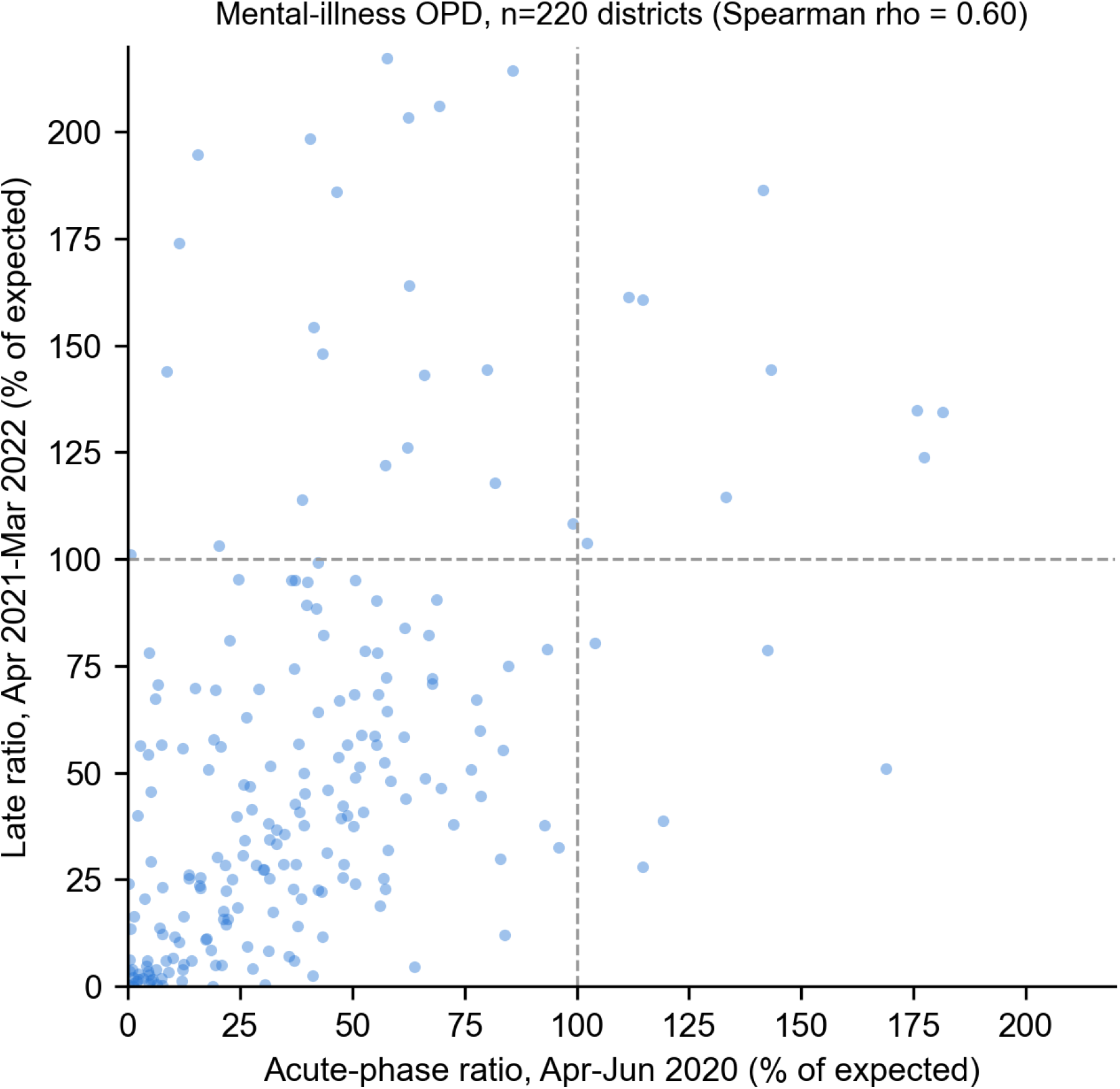
District-level acute shock (April-June 2020) versus final-year recovery (April 2021-March 2022), mental-illness OPD, n=220 districts with stable reporting.

## Discussion

Across five years of routine administrative data spanning both major COVID-19 waves, public mental-health outpatient care in India emerges as the service line that was hit hardest and healed least. Three quantitative facts anchor this conclusion: an acute collapse to 31% of expected in April 2020, deeper than any maternal-child indicator in the same system; a final-year recovery to only 58% of expected — roughly half the recovery achieved by physical-health services measured identically; and 2.2-8.4 million cumulative missed outpatient contacts over two years, against the backdrop of a pre-existing treatment gap already among the world’s largest [2].

These findings convert what has until now been a survey-based impression [3-5] into a national, quantitative, district-resolved estimate, and they carry three implications. First, the mental-physical recovery gap suggests that supply-side restoration alone (reopening facilities, restoring staff) was not sufficient for mental healthcare, whose utilisation depends more heavily on outreach, follow-up continuity, and destigmatised access — all disproportionately damaged by distancing measures and the redeployment of DMHP staff to pandemic duties. Second, the pronounced district scarring argues for targeted restoration: the districts that lost most remained furthest behind, so uniform national programming will systematically under-serve precisely the areas with the largest accumulated unmet need. Third, our end-of-window figures form the natural baseline for evaluating the National Tele Mental Health Programme (Tele-MANAS), launched in October 2022, which has since logged call volumes rising from roughly 12,000 per month in late 2022 to over 90,000 by mid-2024 [6]: whether that channel substitutes for, or merely supplements, the missing facility-based care documented here is a key evaluation question our estimates make tractable.

An alternative explanation — that the deficit reflects deteriorated reporting rather than deteriorated service delivery — cannot be fully excluded, but is unlikely to account for the pattern: physical-service indicators from the same facilities, reported through the same forms in the same months, recovered to near-baseline, so a pure reporting artefact would have to be selective for mental-health line items. Some genuine substitution toward teleconsultation (eSanjeevani) and private providers also likely occurred and is invisible to HMIS; however, the scale of documented teleconsultation growth in 2020-21 was far too small to offset millions of missed public OPD contacts, and private substitution is implausible for the low-income populations DMHP primarily serves.

### Strengths and limitations

Strengths include national coverage at monthly resolution across both waves, an internal physical-services comparator measured with the identical model and reporting system, dual counterfactuals bounding all deficit estimates, and district-level resolution with explicit stability filtering. Limitations: HMIS measures public-facility attendance, not need, diagnosis-verified caseload, or private-sector activity; the mental-illness OPD item is a single aggregate without diagnostic breakdown; district reporting of this item is sparser and noisier than for maternal-child indicators (only 304 districts supported model fitting, and 84 were excluded for instability, limiting generalisability of district-level estimates); and the steep pre-pandemic growth trend makes the trend-based counterfactual generous, which is why we emphasise the range down to the conservative no-trend bound. Finally, ecological service counts cannot identify individual-level consequences of missed care.

## Conclusion

COVID-19 did not merely interrupt India’s public mental healthcare; it set it back years, and — unlike physical-health services — it had not recovered by March 2022. District-targeted restoration and rigorous evaluation of Tele-MANAS against the baseline quantified here should be treated as priorities of the National Mental Health Programme.

## Data Availability

All data produced in the present study are available upon reasonable request to the authors.

https://hmis.mohfw.gov.in

